# The ferroxidase HEPHaestin in Lung cancer: Pathological Significance and Prognostic Value

**DOI:** 10.1101/2021.01.22.21250298

**Authors:** Paola Zacchi, Beatrice Belmonte, Alessandro Mangogna, Gaia Morello, Letizia Scola, Anna Martorana, Violetta Borelli

## Abstract

Iron is a fundamental nutrient utilized by living cells to support several key cellular processes. Despite its paramount role to sustain cell survival, excess of labile iron availability can inflict severe cell damage via reactive oxygen species generation which, in turn, can promote neoplastic transformation. The lung is particularly sensitive to iron-induced oxidative stress, given the high oxygen tensions herein present. Moreover, cigarette smoke as well as air pollution particulate can function as vehicles of iron supply, leading to an iron dysregulation condition shown to be crucial in the pathogenesis of several respiratory diseases including lung cancer. Hephaestin (HEPH) belongs to a group of exocytoplasmic ferroxidases emerged to contribute to cellular iron homeostasis by favouring its export. Although HEPH can affect the concentration of intracellular iron labile pool, its expression in lung cancer and its influence on prognosis have not been investigated.

In this study we explored the expression pattern and prognostic value of HEPH in the most prevalent histotypes of lung cancers including lung adenocarcinoma and lung squamous cell carcinoma across in silico analyses using UALCAN, Gepia and Kaplan-Meier plotter bioninformatics. We took advantage of TIMER to assess the correlation between HEPH and tumour infiltrating immune and non-immune cells. Then we performed immunohistochemical analysis to dissect the presence of HEPH either in “healthy” and tumor lung tissues. Overall, our data suggest a positive correlation between higher level of HEPH expression with a favorable prognosis in both cancer histotypes.

## Introduction

Lung cancer represents the most frequent malignant neoplasm in most countries and the leading cause of death worldwide in both sexes (1). The incidence of lung cancer is low in people aged below 40 years but it dramatically increases up to age 60-65 years in most populations. The most common subtype of lung cancer is non-small cell lung cancer (NSCLC; 85%), classified into lung adenocarcinoma (LUAD), the most prevalent form, followed by lung squamous cell carcinoma (LUSC) and large cell carcinoma (2). Smoking status is certainly the most important causative link in lung cancer development even though air pollution represents another paramount source of risk factor. Airborne Particulate matter (PM), in particular the small size components (PM_10_, PM_2.5_ and ultrafine particles-UFP), which include combustion products, soot, exhaust emission from vehicles and industrial processes, have attracted attention mainly for two reasons: firstly, these particles, due to their small size, remain suspended in the air for quite a long time, thus increasing the chance of being inhaled; secondly, these particles are vehicles of chemical compounds, in particular transition metals, being iron present in significant concentration (3). Iron is also found in cigarette smoke, the strongest causative link to pulmonary pathology.

Iron toxicity relies on its high redox cycling reactivity which can drive the production of free radical species (ROS) known to promote many aspects of tumour development and progression (4). The lung is extremely sensitive to metal-induced oxidative stress due to its unique role for massive oxygen transfer into the bloodstream (5). Therefore, as a protective strategy to prevent ROS generation, lung epithelial cells have evolved a tight control on iron import, storage and export aimed at keeping its absolute concentration low, while sustaining the metabolic demand (6 REF Ghio 2009). Efficient iron uptake and intracellular sequestration can limit its toxicity but long-term storage can increase the probability of its possible mobilization, resulting in oxidative cell damage. Iron export mechanisms are therefore necessary to prevent its excessive intracellular accumulation, as it may occur upon exogenous iron supply via airborne pollutants inhalation. The only known non-heme iron export pathway relies on the activity of the transmembrane ferrous iron transporter ferroportin 1 (FPN1) also known as solute carrier family 40 member 1 (SLC40A1) (7) in conjunction with the large membrane–anchored copper-dependent ferroxidase (FOX) Hephaestin (HEPH), required to oxidized iron to its ferric form (8). In enterocytes FPN1 allows the translocation of iron across the basolateral membrane and its release into the bloodstream (9). In lung FPN1 is, instead, mainly expressed in the apical membrane of the airway epithelium (10) where it is supposed to promote iron release into the airways or the lumen of the alveoli to meet the need of detoxification. This egress pathway has been shown to be compromised in diverse types of cancers (11). In particular FPN1 mRNA expression levels appeared significantly down-regulated in lung tumour as compared to matched healthy tissue, a condition that is expected to guarantee an increase in the intracellular labile iron pool necessary for all metabolic processes involved in cell proliferation (12). The role played by HEPH in iron metabolism in lung is still poorly characterized as well as its possible contribution to lung carcinogenesis and growth. We recently identified a single-nucleotide polymorphism within HEPH gene, leading to a missense variation of this multicopper ferroxidase, which results protective against asbestos-dependent malignant pleural mesothelioma and lung carcinoma (13, 14). Moreover, in breast cancer HEPH expression has been shown to be down-regulated by the histone methyltransferase G9a, leading to changes in iron homeostasis that burst cancer growth (15).

In the current study, we examined the expression and prognostic value of HEPH expression in LUAD and LUSC patients in databases such as UALCAN, GEPIA and Kaplan-Meier plotter. Moreover, we investigated the correlation of HEPH expression with tumour-infiltrating immune and non-immune cells characterizing the tumour microenvironment via Tumour Immune Estimation Resource (TIMER). Finally, we evaluated the distribution of endogenous HEPH in lung cancer tissues. These data altogether further support the key role played by iron dysregulation in the carcinogenic mechanism of lung malignancies

## Materials and Methods

### Gene expression and Survival analysis

Our analysis focused on the prognostic effect of *HEPH* gene in lung adenocarcinoma (LUAD) and in lung squamous cell carcinoma (LUSC). The expression level of the gene in different carcinomas was analyzed using UALCAN (http://ualcan.path.uab.edu) and GEPIA (http://gepia.cancer-pku.cn). Those tools estimate the effect of gene expression level on the patient survival in addition to be a web resource for analyzing cancer transcriptome data (16, 17). We compared the differences in mRNA level between cancers and normal tissue, using genomics data from “The Cancer Genome Atlas” (TCGA lung). The prognostic significance of *HEPH* mRNA expression and survival in LUAD and LUSC were analyzed by Kaplan Meier plotter (https://kmplot.com/analysis). Kaplan Meier plotter use genomics data from Gene Expression Omnibus, and European Genome-phenome Archive to generate survival probability plots and to perform survival analysis (2, 3). The same analysis was done for other genes (*ACTA2, FAP, PDGFRA, PDGFRB, PECAM1, vWF*). The hazard ratio with 95% confidence intervals and log-rank *p*-value were also computed..

### Protein Expression Analysis

The expression HEPH proteins between cancer and normal tissue were analyzed using UALCAN, which provided protein expression analysis option using data from Clinical Proteomic Tumor Analysis Consortium (CPTAC) Confirmatory/Discovery dataset (4). Unfortunately, at the time of writing, UALCAN tool provided data only for LUAD histotype.

### TIMER Database Analysis

TIMER is a comprehensive resource for systematic analysis of immune infiltrates across diverse cancer types (www.cistrome.shinyapps.io/timer/) (18). TIMER applies a statistical method to infer the abundance of tumor-infiltrating immune cells (TIICs) from gene expression profiles using data from the TCGA dataset (19). We analyzed *HEPH* expression in lung cancers and the correlation between its expression with the abundance of immune infiltrates, including B cells, CD4+ T cells, CD8+ T cells, neutrophils, macrophages, cancer associated fibroblasts and endothelial cells via gene modules. These gene markers are referenced in prior studies (7, 8). Gene expression levels against tumor purity are also displayed (20, 21). The correlation module generated the expression scatter plots between several genes and defined genes of TIICs in chosen carcinomas, together with the Spearman’s correlation and the estimated statistical significance. Several genes were used for the x-axis, and related marker genes are represented on the y-axis as genes of TIICs. The gene expression level was displayed with log2 RSEM.

### Statistical Analysis

Survival curves were generated by the Kaplan Meier plotter (22). All results are displayed with *p*-values from a log-rank test. *p*-values < 0.05 were considered significant. In TIMER, the correlation of gene expression was evaluated by Spearman’s correlation and statistical significance, and the strength of the correlation was determined using the following guide for the absolute value: 0.00 - 0.19 “very weak,” 0.20 - 0.39 “weak,” 0.40 - 0.59 “moderate,” 0.60 - 0.79 “strong,” 0.80 - 1.0 “very strong”.

### Immunohistochemistry analysis on tumour tissues

All tissue samples of lung adenocarcinoma and lung squamous cell carcinoma enrolled for this study were collected according to the Helsinki Declaration and the study was approved by the University of Palermo Ethical Review Board (approval number 09/2018). Surgical normal tissue samples of lung and the malignant counterpart were selected for immunohistochemical analysis for HEPH expression. Invasive malignant neoplasia specimens included the two more represented histotypes including LUAD and LUCS. The study was approved by the Institutional review board of the University of Palermo (09/2018). A specific informed consent was not required at the time of tissue sample collection for the immunohistochemical analysis of archival tissue sections since the patients were not identified and genetic analysis was not carried out.

Immunohistochemistry was carried out on FFPE human tissue sections. Briefly, sections 4 micron-thick were cut from paraffin blocks, dried, de-waxed and rehydrated. The antigen unmasking technique was performed using Target Retrieval Solutions, pH=9 EDTA-based buffer in thermostatic bath at 98°C for 30 minutes. After the sections were brought at room temperature, the neutralization of the endogenous peroxidase with 3% H_2_O_2_ and protein blocking by a specific protein block were performed. For HEPH immunostaining, sections were probed with mouse monoclonal anti-human HEPH (Diluition 1:100, pH 6, Clone sc-365365 Santa Cruz Biotecnology) overnight at 4°C. Antibody-Antigen recognition was detected using Novolink Polymer Detection Systems (Novocastra Leica Biosystems, Newcastle) and employing the high sensitivity AEC (3-Amino9-Ethylcarbazole) as chromogen. Slides were counterstained with Harris Haematoxylin (Novocastra, Ltd).

All the sections were analyzed under Zeiss Axio Scope A1 optical microscope (Zeiss, Germany) and microphotographs were collected using an Axiocam 503 Color digital camera with the ZEN2 imaging software (Zeiss Germany).

## RESULTS

### The mRNA Expression Levels of HEPH in Different Types of Human Cancers

The ferroxidase HEPH has recently emerged to play a role in breast tumour cell growth; in particular its decreased expression has been significantly correlated with a poor survival in affected patients (15). In order to expand the analysis to other cancer types we examined HEPH expression using UALCAN that analyse TCGA RNA-sequencing and patients’ clinical data from 33 different cancer types, including several metastatic tumors (22). This analysis unveiled that a significant down-regulation of the HEPH mRNA expression levels is found in several other malignancies such as BLCA (bladder urothelial carcinoma), BRCA (breast invasive carcinoma), COAD (colon adenocarcinoma), KICH (kidney chromophobe), KIRP (kidney renal clear cell carcinoma), LIHC (liver hepatocellular carcinoma), LUAD (lung adenocarcinoma), LUSC (lung squamous adenocarcinoma), PRAD (prostate adenocarcinoma), READ (rectum adenocarcinoma), and UCEC (uterine corpus endometrial carcinoma) compared to the corresponding normal tissues (Fig. 1A).

**Figure 1.**
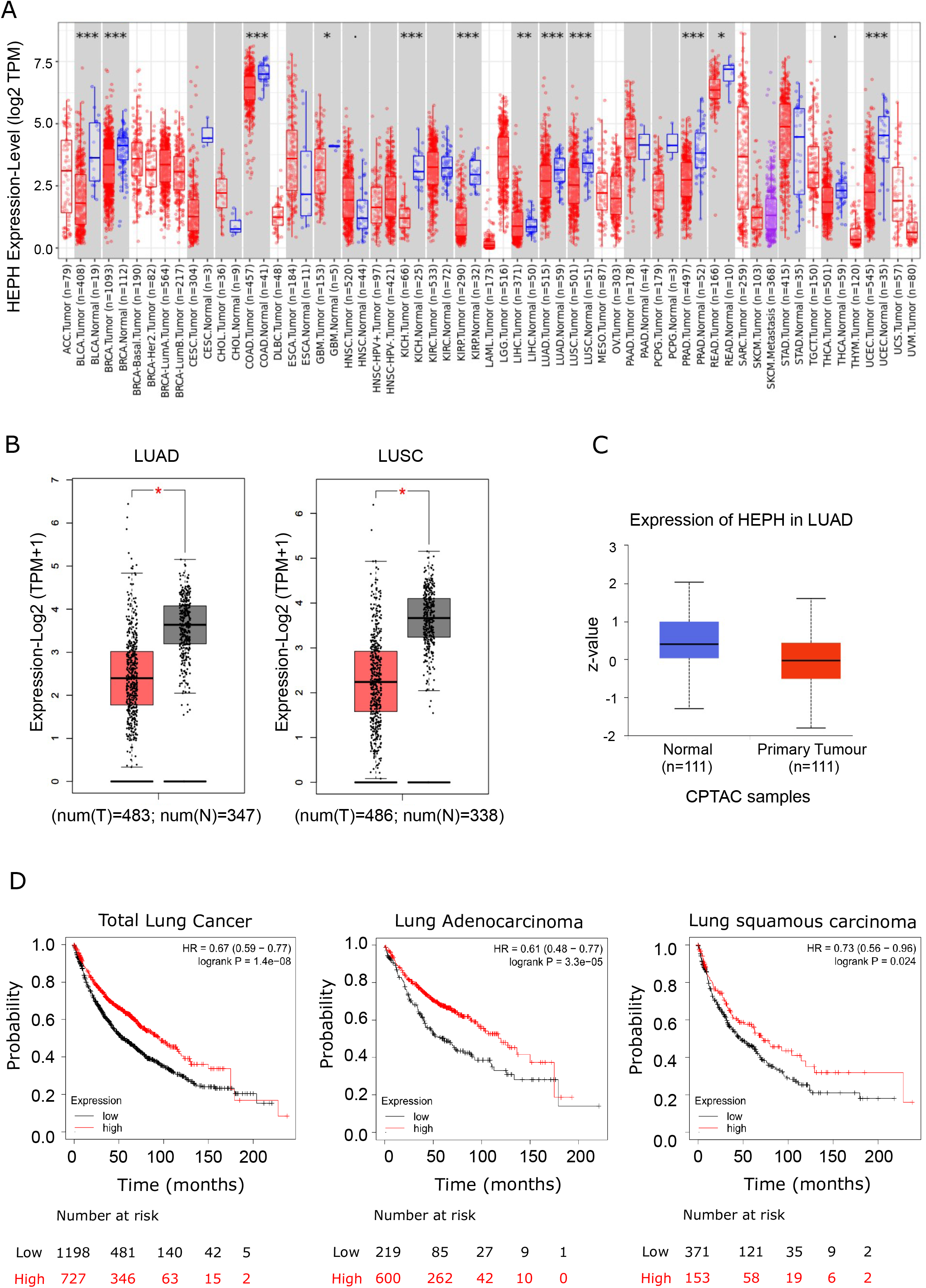
Pathological significance of HEPH expression in different types of human cancers and in-depth evaluation in LUAD and LUSC. (A) Human HEPH expression levels in different tumour types from TCGA database were determined by TIMER (*P < 0.05, **P <0.01, ***P < 0.001). (B) HEPH mRNA expression comparisons between normal and tumour tissues were further obtained from the GEPIA web tool. (C) HEPH protein expression comparison between normal and tumour tissues obtained from the UALCAN web tool (Wilcoxon test). P-value < 0.05 was used to assess differences.(D) Survival analyses of HEPH by Kaplan–Meier estimator with log-rank test obtained from the Kaplan Meier plotter web tool. Survival differences are compared between patients with high and low (grouped according to Auto select best cut-off) expression of HEPH. H, high expression; L, low expression.

Given our interest in better understanding the role iron’s dysregulation may exert in lung cancer development and prognosis, we evaluated HEPH mRNA expression levels in the most prevalent histological types LUAD and LUSC, as compared to normal tissue, utilizing the GEPIA database. Consistently with the previous analysis, a significant decrease in HEPH mRNA expression was found in LUAD and LUSC, as compared to healthy controls (Fig. 1B) and this reduction was confirmed at the protein level only for LUAD histotype, based on UALCAN dataset (Fig. 1C), since corresponding proteomic data for LUSC are still not available.

To investigate the correlation between HEPH expression and patients’outcome we took advantage of the Kaplan-Meier overall survival curves to establish and compare the survival differences between patients with high and low expression of the ferroxidase (grouped according “Auto select best cutoff”) (Fig. 1D). In both LUAD and LUSC datasets, the high expression group had a significantly longer overall survival than the low expression group, thus indicating that higher HEPH expression correlates with a better prognosis.

### HEPH expression is correlated with immune and non-immune infiltration

It is well established that cancer cells are characterized by an iron-seeking phenotype, which is fundamental to support the enhanced metabolic demand characteristic of actively proliferating cells (23). This increased request of iron supply is achieved not only upon up-regulating iron import pathways while down-regulating storage and export routes but also by altering how other cell types of the tumor microenvironment, including immune cells, endothelial cells, pericytes and fibroblasts, metabolize iron. We therefore assessed the correlations of HEPH expression with immune and non-immune infiltration levels using TIMER web resource (22). In particular, we assessed, as immune infiltrate, B cells, CD4+ T cells, CD8+ T cells, macrophages and dendritic cells while cancer associated fibroblasts and endothelial cells were analysed as infiltrating non-immune cell types. The results showed that HEPH expression had a significant negative correlation with tumour purity, the parameter identifying the proportion of cancer cells present in the tumour tissue, in both type of lung cancers (Figure 2). In addition, HEPH expression demonstrated a very weak correlation with all infiltrating immune elements tested (Table 1), while a strong positive correlation was found only with cancer associated fibroblasts (CAFs) and endothelial cells (ECs) (Figure 2).

**Table 1.**
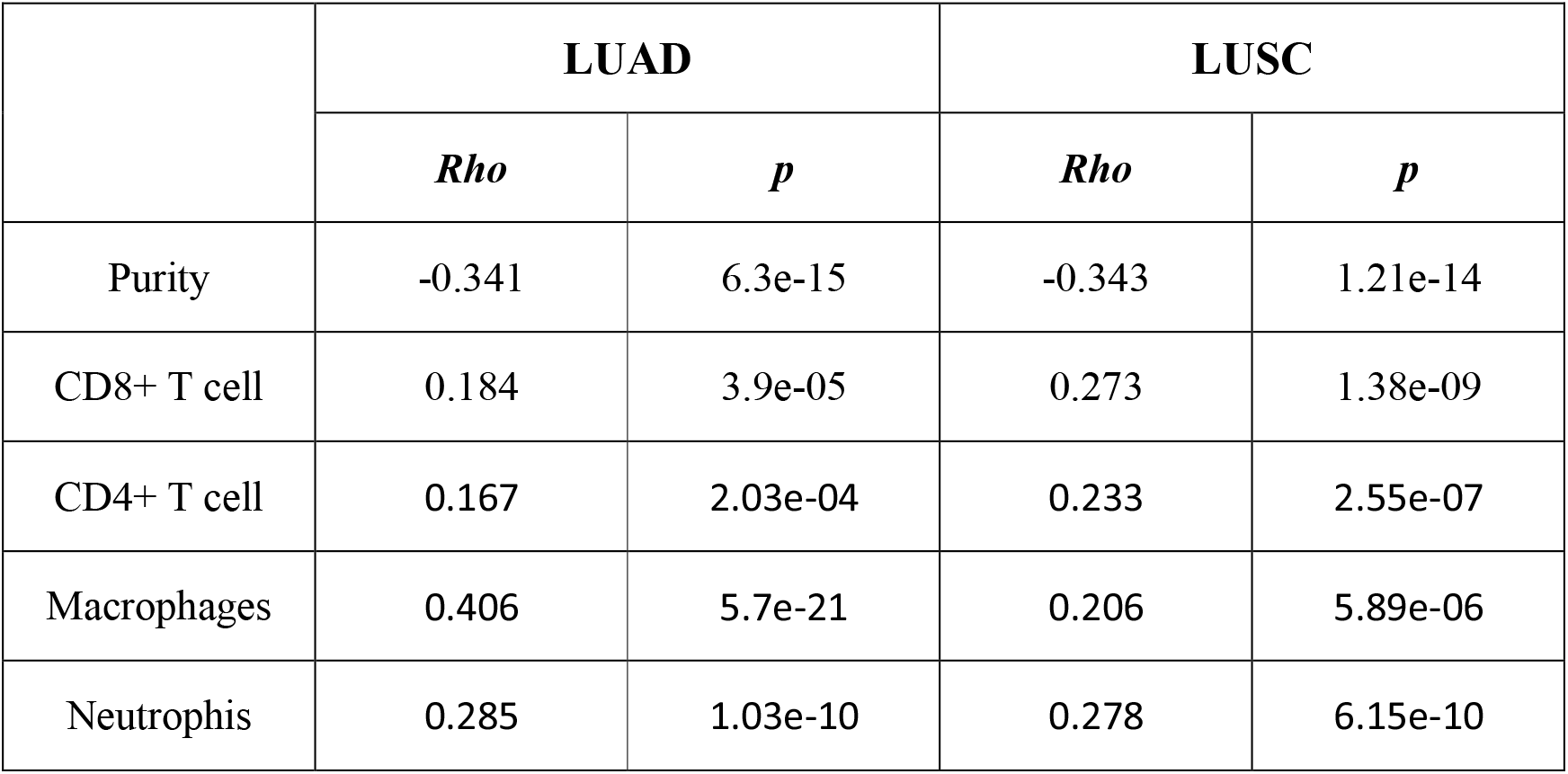
Correlation analysis between HEPH expression and immune infiltration level of the indicated immune cells. The “Purity Adjustment” option was applied to all analysis performed.

|  | LUAD |  | LUSC |  |
| --- | --- | --- | --- | --- |
|  | *Rho* | *p* | *Rho* | *p* |
| Purity | -0.341 | 6.3e-15 | -0.343 | 1.21e-14 |
| CD8+ T cell | 0.184 | 3.9e-05 | 0.273 | 1.38e-09 |
| CD4+ T cell | 0.167 | 2.03e-04 | 0.233 | 2.55e-07 |
| Macrophages | 0.406 | 5.7e-21 | 0.206 | 5.89e-06 |
| Neutrophis | 0.285 | 1.03e-10 | 0.278 | 6.15e-10 |

**Figure 2.**
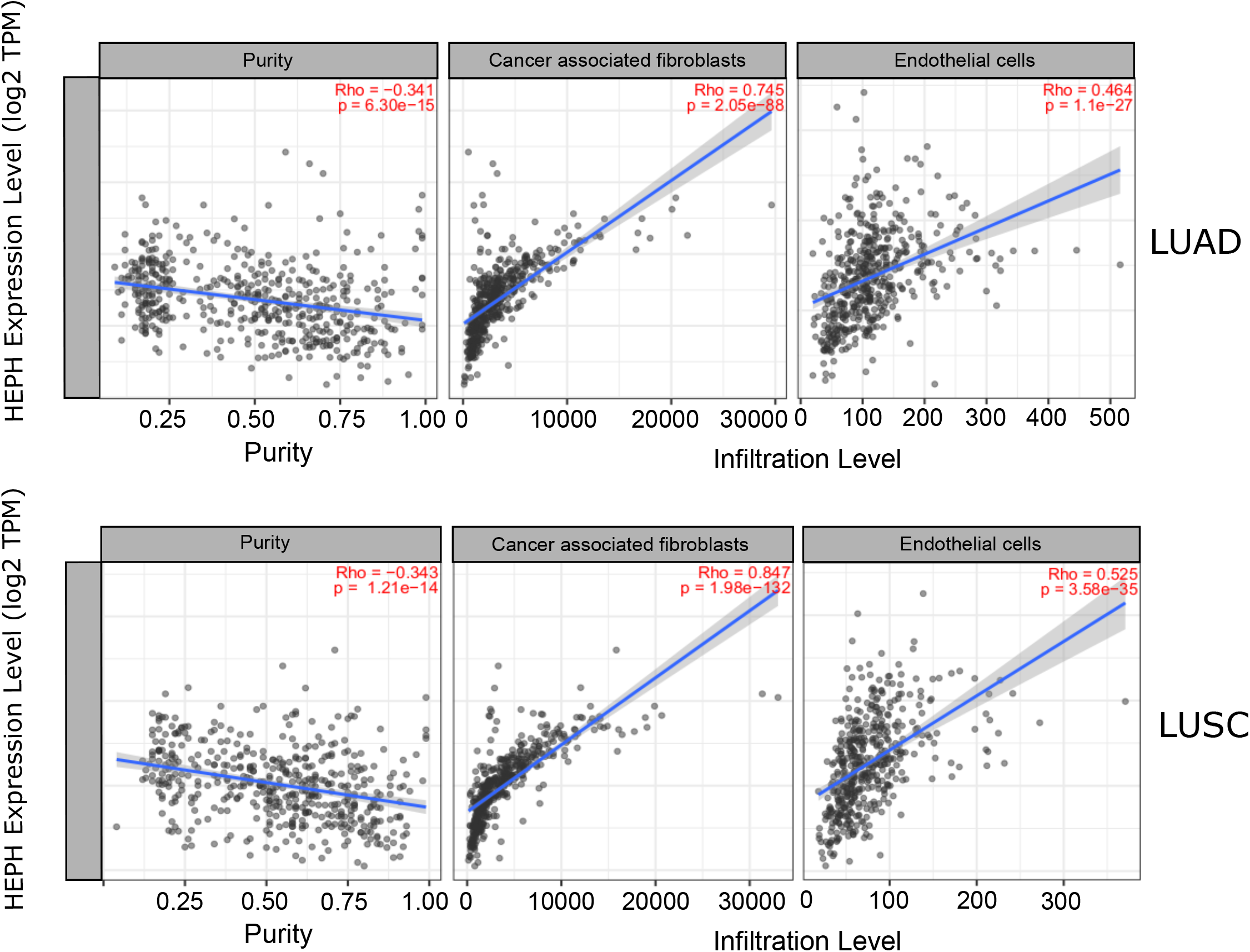
Correlation of HEPH expression with infiltration level of non-immune cells in LUAD and LUSC. HEPH expression is significantly negatively related to tumour purity and has significant positive correlations with infiltrating levels of cancer-associated fibroblasts and endothelial cells.

CAFs are the most abundant cells in solid cancer. They can derived from several sources including activation of resident fibroblasts (24), epithelial-mesenchymal transition of epithelial cells (25), endothelial-mesenchymal transition of resident endothelial cells (26). Compared to normal fibroblasts they are characterized by enhanced proliferative and migratory features, and they are also more metabolically active. Tumour endothelial cells refers to the cells lining the tumour-associated blood vessels that provide nutrition and oxygen to the tumour, contributing to its growth and development. They also constitute one of the main sources of cancer-associated fibroblasts (CAFs).

To further characterized the relationship between HEPH and these infiltrating cells in lung malignancies we explore the correlation between HEPH and a list of marker sets known to be widely used to identified CAFs and ECs, using the TIMER Gene Correlation module. In particular we used α-SMA (ACTA2, marker also for vascular muscular cells and pericytes), fibroblasts activation protein (FAP, also expressed in a subset of CD45+ immune cells), plateled-derived growth factor receptor-α/β (PDGFRA/B) as biological markers for CAFs (Figure 3A); PECAM1 (CD31) and von Willebrand Factor (vWF) as markers for the endothelial cells (Figure 3B). After the correlation adjustment by tumour purity, HEPH expression level was significantly correlated with all marker sets tested (Figure 3).

**Figure 3.**
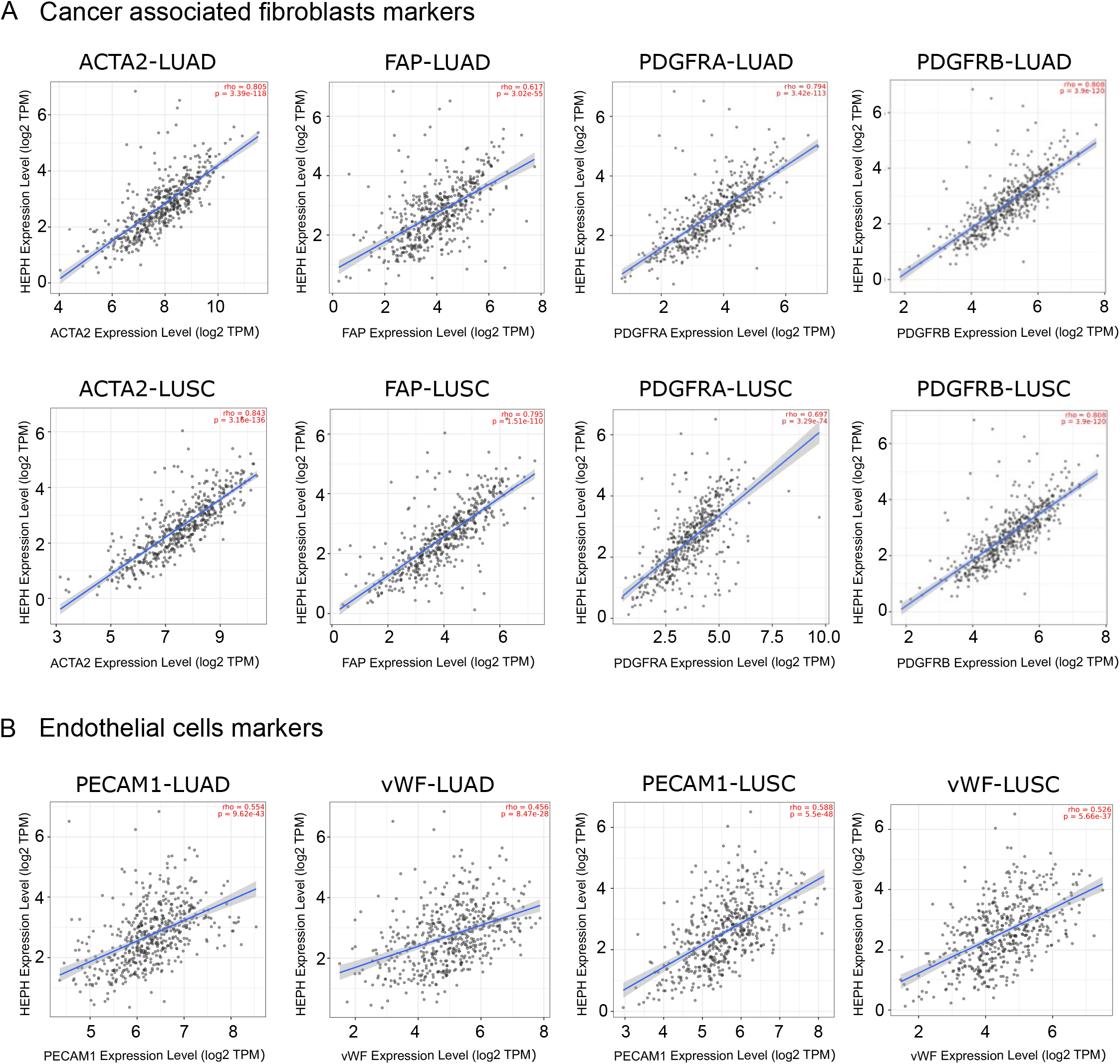
HEPH expression positively correlated with markers of cancer-associated fibroblasts and endothelial cells in both LUAD and LUSC. Scatterplots of correlations between HEPH and gene markers include ACTA2, FAP, PDGFRA, PDGFRB for cancer-associated fibroblasts and PECAM1 (CD31) and vWF for endothelial cells.

Interestingly we also found that the mRNA expression level of all these marker genes, with the only exception of FAP, were significantly down-regulated in both lung malignancies as compared with paired normal tissues, based on GEPIA datasets (Figure 4A). Moreover, Kaplan Meier analysis indicated that high expression of ACTA2 and PDGFRA as well as PECAM1 and vWF were associated with better overall survival, as it is for HEPH expression (Figure 4B).

**Figure 4.**
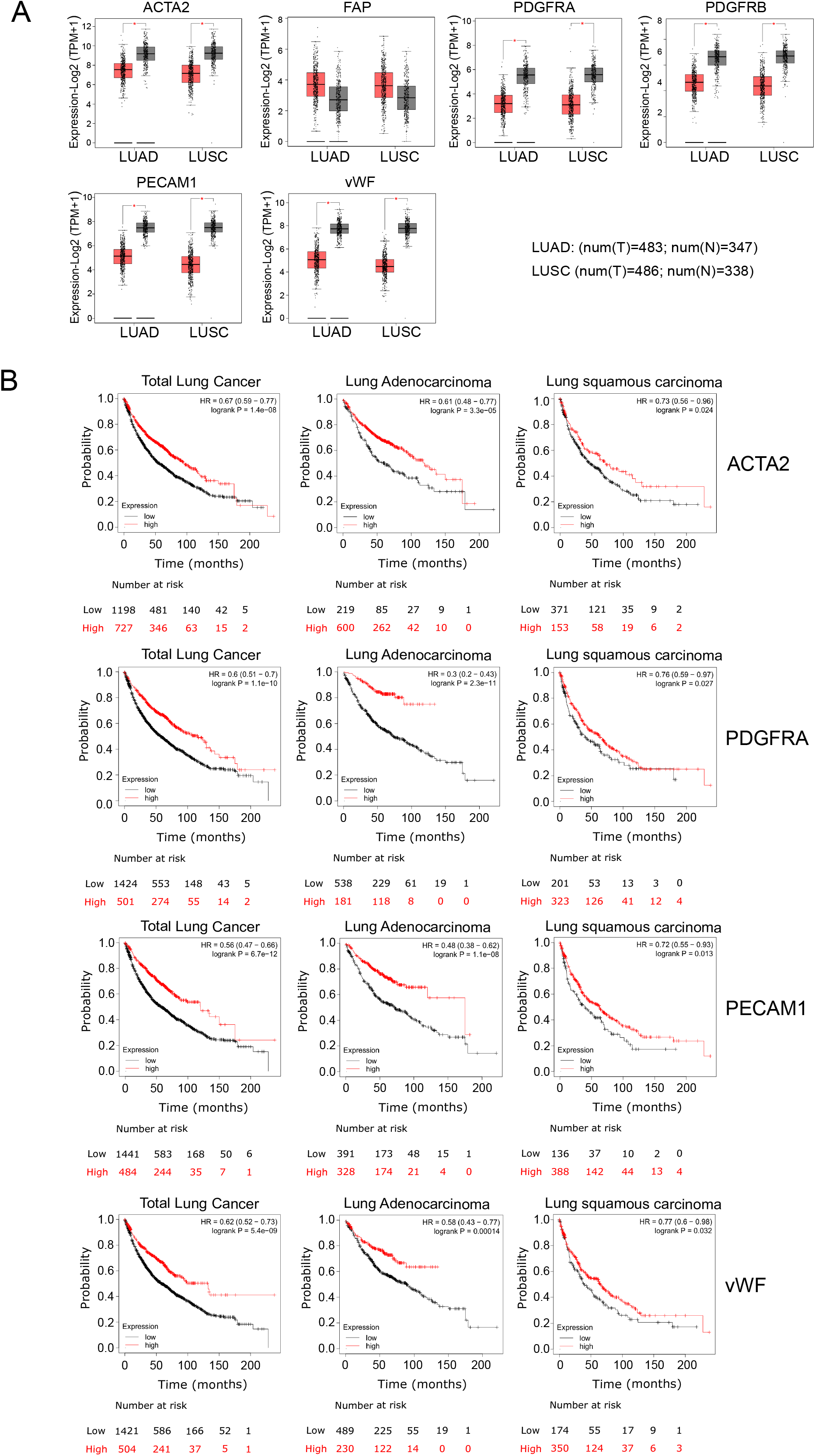
Overall survival curve of each cancer-associated fibroblasts and endothelial marker shown to correlate to HEPH expression and produced by Kaplan-Meier website resource. OS differences are compared between patients with high and low (grouped according to Auto select best cut-off). H, high expression; L, low expression.

### Distribution of HEPH in clinical LUAD and LUSC specimens

Based on the results obtained upon TIMER database analysis we aimed at better understanding the distribution of the HEPH in a series of specimens of LUAD and LUSC upon ferroxidase immunohistochemical labeling. As shown in Figure 5 panel A and B, the ferroxidase staining was quite intense on endothelial cells surrounding the vasculature in peri-tumour tissues, being endothelial cells and possibly perycites immunoreactive for HEPH primary antibody. HEPH was also associated to macrophages, as identified by their spherical appearance and by the presence of a reniform nucleus, and neutrophils based on their small round shape and on a clearly identifiable multi-lobed nucleus (Figure 5B, see arrows).

**Figure 5.**
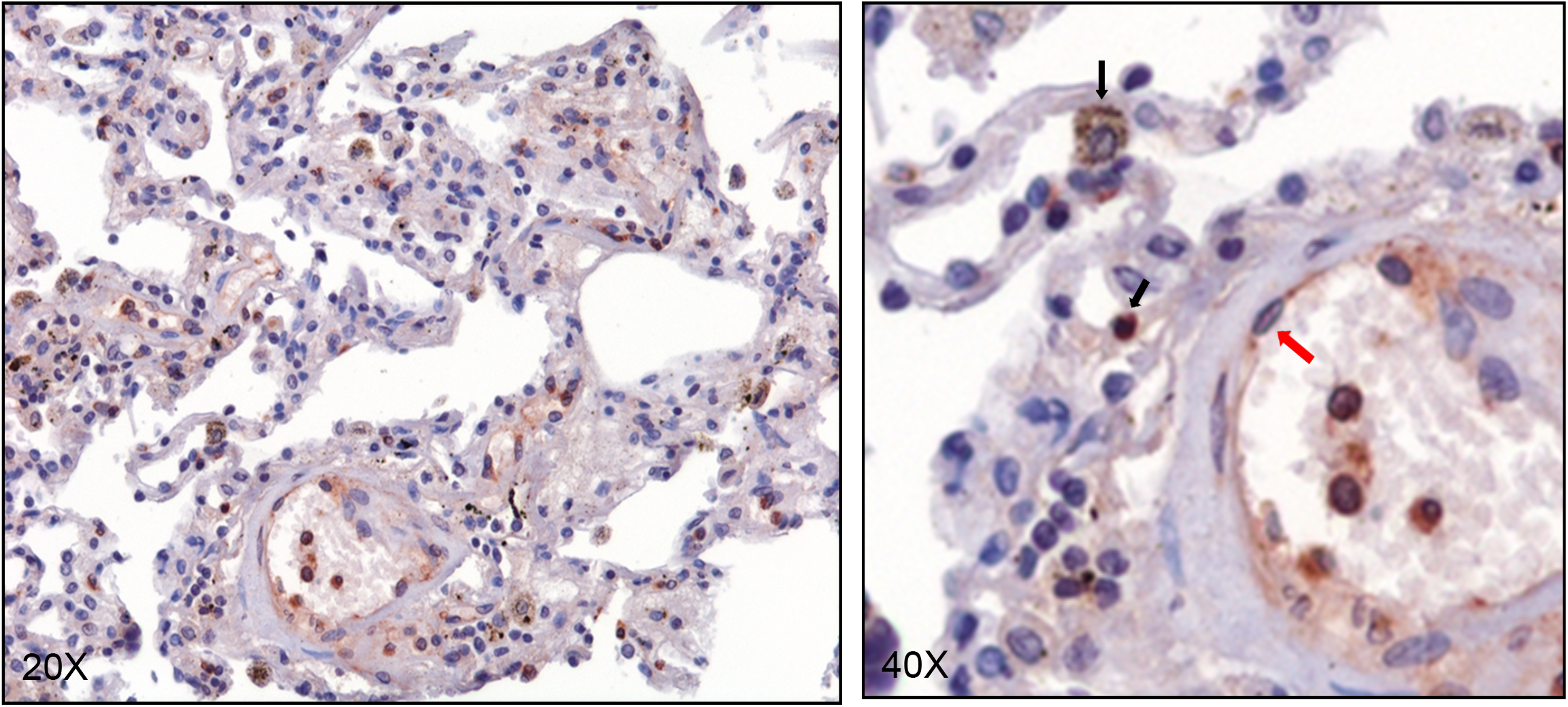
HEPH distribution in “normal” lung. Representative microphotographs relative to HEPH expression by the endothelial cells (red arrows), cells of the myelomonocyte lineage cells including monocytes, macrophages, and granulocytes (black arrows) in “healthy” pulmonary parenchyma. Original Magnification 20X (A) and 40X (B).

In cancer tissues HEPH distribution appeared quite different in the two malignancies analyzed. In particular, in LUAD specimens we noticed that tumoral cells were totally lacking HEPH expression while the very few cells HEPH positive infiltrating the cancerous tissue were characterized by either a neutrophil-like or a spindle-like fibroblast morphology (Figure 6, panels B and D). There were also some macrophages immune-labeled, identified based on their morphological features. In LUSC, instead, we observed that about 30% of cancer cells, identified for their characteristic large polygonal shape, were positive, to variable extent, for HEPH labeling (Figure 7, panel B). Moreover, the tumour mass was massively infiltrated by HEPH-expressing cellular elements of neutrophil-like morphology and possibly also some monocyte/macrophages (Figure 7, panels A and C). There were also a consistent amount of HEPH positive mesenchymal cells exhibiting spindle-shaped morphology and recognized as fibroblastic stromal component (Figure 7, panels D).

**Figure 6.**
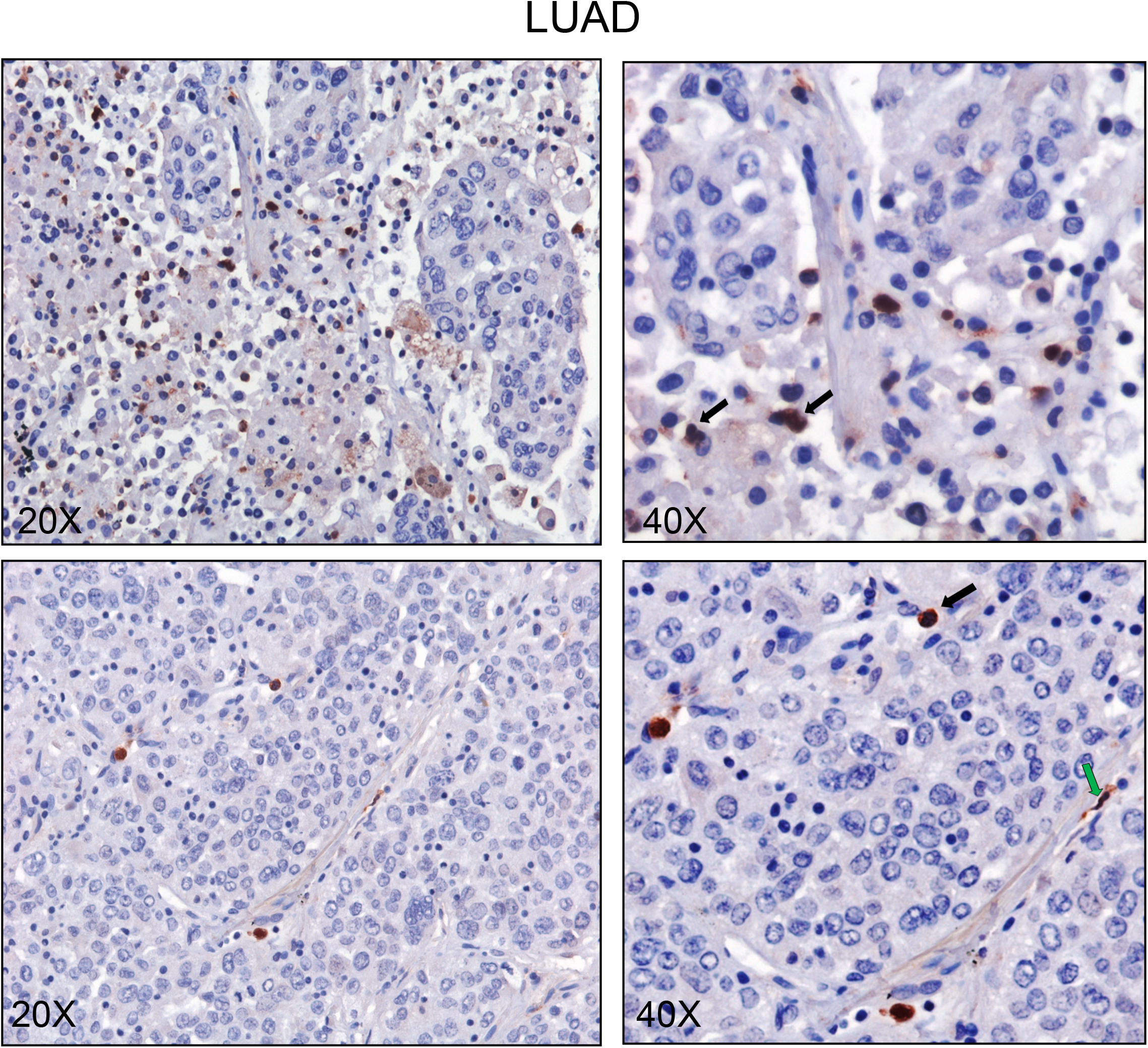
HEPH distribution in lung adenocarcinoma specimen. Representative microphotographs relative to HEPH expression in two different lung adenocarcinoma affected patients (A-C and higher magnification B-D). The tumour tissue is infiltrated by some HEPH positive myelomonocyte lineage cells including monocytes, macrophages, and granulocytes (black arrows). Original Magnification 20X (A) and 40X (B).

**Figure 7.**
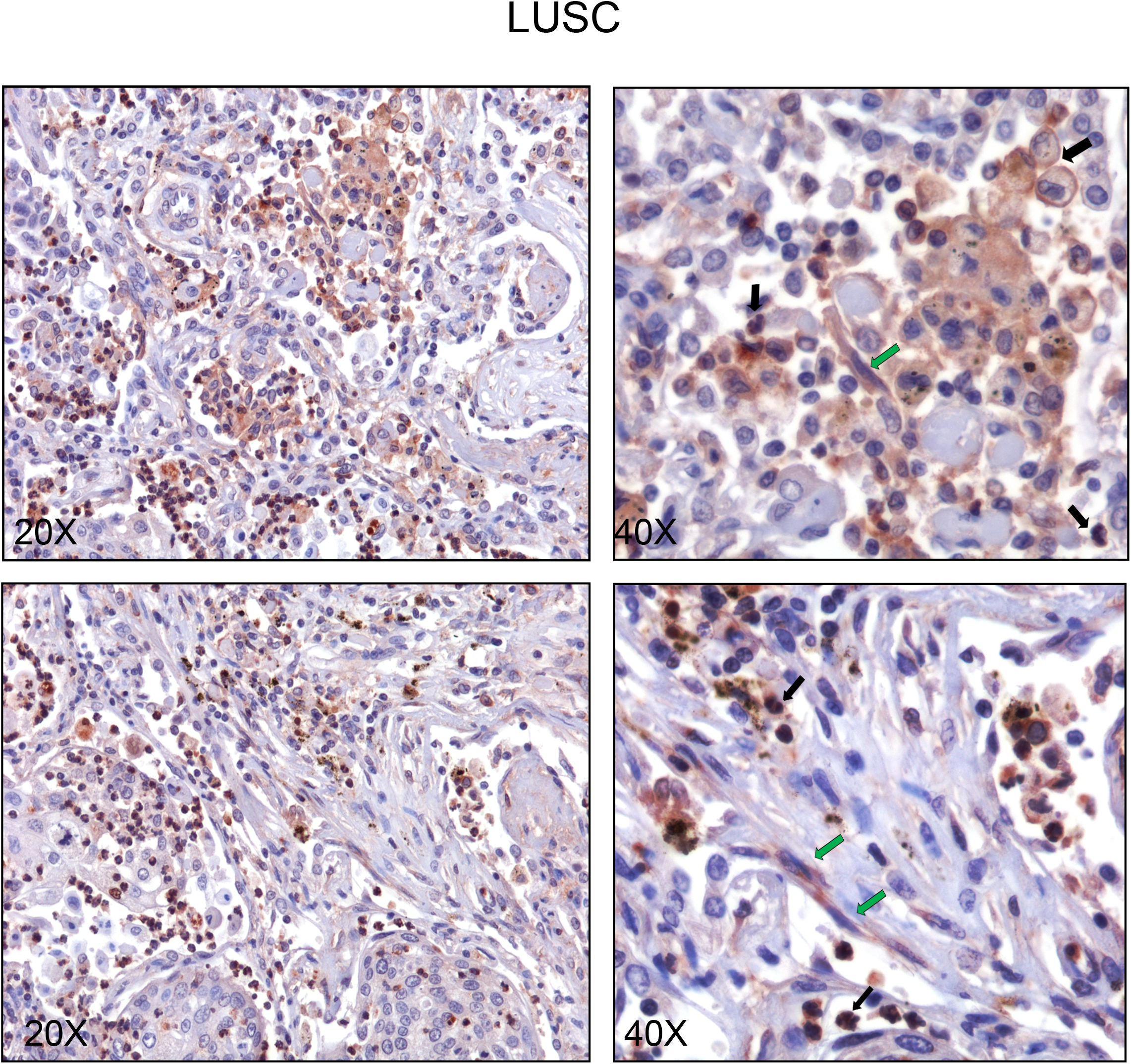
HEPH distribution in lung squamous cell carcinoma specimen. Representative microphotographs relative to HEPH expression in two different lung squamous cell carcinoma affected patients (A-C and higher magnification B-D). The tumour tissue is infiltrated by some HEPH positive stromal elements (panel D, green arrows) and cells of the myelomonocyte lineage cells including monocytes, macrophages, and granulocytes (black arrows). Original Magnification 20X and 40X (B).

These immune-labeling experiments, in agreement with the bioinformatics analysis, confirm the expression of HEPH by stromal elements infiltrating the tumor microenvironment and endothelial cells; in addition we clearly detected expression of the ferroxidase in cellular elements belonging to the innate-immunity, such as macrophages and neutrophils, as identified based on morphological features.

## Discussion

Lung cancer still represent the leading cause of cancer-related deaths both in men and women, especially in developed countries. Lung adenocarcinoma is the most common histologic subtype, whose incidence has dramatically arose and overcome that of squamous cell carcinoma due to an increase in the incidence of lung cancer in women. Despite advances in diagnosis and treatments, the overall 5-year survival rate remains dismal especially when lung cancer is diagnosed at advanced stages (27). Therefore, a better understanding of the molecular mechanisms underlying lung carcinogenesis should contribute to the development of novel strategies for its preventions and therapy. Cigarette smoking represents the principal risk factor for lung cancer, but inhalation of air pollutants is also accountable for the increased incidence of this type of malignancies. Air pollution and tobacco smoking have been shown to impact on lung iron metabolism, being the source of additional iron supply on a tissue which is physiologically exposed to oxidative stress. In the present studies we identified HEPH, a protein involved in exporting iron out from the cell, as promising predictor for the clinical prognosis in lung cancer.

HEPH is a multi-copper oxidase whose function has been better characterized in small intestine, where it is required for iron egress from the enterocyte into the circulatory system. HEPH is thought to work in concert with Ferroportin (FPN1), the only known mammalian iron exporter for non-heme iron, whose down-regulation has been monitored in several cancers and it is usually correlated to a poor prognosis (12). FPN1 reduction is thought to act by increasing the concentration of the intracellular labile iron pool, the catalytic/reactive iron, which, on one side, is required to sustain the high metabolic demand of actively proliferating cells, but, on the other hand, it can trigger the production of free radical species, further boosting the oxidative damage. In lung FPN1 is facing the lumen of the alveoli and this localization has been attributed to a role in iron detoxification (10). Indeed, environmental iron arriving to the lung epithelium can be initially buffered by the activity of the antioxidant molecules such as ascorbic acid, reduced glutathione, and mucin, and once loaded on transferrin and lactoferrin herein present, it can undergo transferrin receptor 1 (TfR1) and lactoferrin receptor (LfR) internalization by epithelial alveolar cells and alveolar macrophages and be safely stored bound to ferritin (28). Under iron overload condition the excess of pulmonary iron can be release into the lumen of the alveoli via FPN1 permease and possibly oxidize by GPI-anchored or soluble ceruloplasmin, an HEPH homologue ferroxidase (29). Based on our immunohistochemical localization studies, HEPH has emerged to be highly expressed not in the lung epithelium, as functional partner of apically located FPN1, but on the endothelial cells of the lung vasculature and in some perivascular cells (perycite/fibroblast cells). HEPH has been already identified on endothelial cells of brain capillaries (30). In this context the ferroxidase has been shown to be localized on the endothelium abluminal side, as well as on perycites, where it is supposed to convert ferrous iron released in the extracellular space by endothelial FPN1 into ferric iron, thus limiting oxidative damage. A similar mechanism could also operate in lung, where endothelial localized HEPH could similarly act on nutritional ferrous iron shipped in the interstitial space by FPN1, making it available for resident cell uptake, thus limiting oxidative damage. This precise architectural organization and functional interplay between endothelial cells, perycites and alveolare epithelial cells is compromised under pathological conditions such as cancer, and somehow reorganize to foster cancer cells proliferation.

Based on our bioinformatics analysis, further corroborated by immunohistochemistry, cancer cells in LUAD and LUSC are mostly not expressing (LUAD) or poorly expressing, with a variable degree, HEPH. This reduction is expected to limit FPN1-dependent iron export, which needs the concomitant ferroxidase activity to fully function as a Fe^2+^ permease. The consequent increase in the intracellular labile iron pool, via ROS generation, can stimulate cell proliferation and survival by inducing several pro-survival signalling pathways and promote epithelial-to-mesenchymal transition (EMT), an essential step for the initiation of metastasis. Innate immune cells and cancer-associated fibroblasts are also a major source of iron and ROS in the tumour microenvironment (31).

According to TIMER bioinformatics analysis, HEPH expression has emerged to be strongly and positively correlated with cancer associated fibroblasts (CAFs). Upon LUAD and LUSC specimens HEPH immunolabeling, we clearly identified HEPH expressing cells characterized by an elongated spindle-shaped morphology typical of fibroblasts, enveloping some tumour nests in LUSC cancer histotype. CAFs are the most dominant cellular component in the tumour stroma which not only provide physical support to tumour cells but also play key role in promoting or hampering tumorigenesis in a context-dependent manner. CAFs are highly heterogenous cell population due to their multiple origins: they can arise from resident fibroblasts, bone marrow-derived progenitor cells or epithelial/endothelial cells that have undergone epithelial to mesenchymal transition. Cellular trans-differentiation, both from stromal cell to stromal cell and from tumour cell to stromal cell have all been described, being fibroblast trans-differentiation into activated myofibroblast, positive for α-SMA expression, the most frequently cited example. With the aim to better characterized our HEPH positive stromal elements we performed HEPH/α-SMA double-labelling experiments but we observe that a restricted minority of cells were for both antigens (data not shown). This observation raises the possibility that these tumour-associated stromal elements could belong to the most aggressive “matrix remodelling” subtypes, as classified by Marini and colleagues (21), which are characterized by increased expression of fibroblast activating protein (FAP) but low expression of α-SMA. It is interesting to note that FAP mRNA expression is the only marker emerged to be slightly upregulated, even though not reaching a statistical significance, in LUAD and LUSC as compared to normal tissue (Figure 4A). Further in-depth studies will be required to unravel all these issues.

As far as innate immunity is concern, neutrophils and macrophages, as detected by MPO and CD14 immunoreactivity, were also identified as HEPH expressing cells infiltrating the tumour tissues under investigation. Neutrophils, the hallmark of acute inflammation, have been shown to represent a substantial proportion of the immune infiltrate in a wide variety of cancer types, including lung cancer, even though their role is still debated (32). Tumour-associated neutrophils (TANs) could exert a beneficial anti-tumour function, based on their high cytotoxic potential that could be directed toward cancer cells (33). On the other hand, other studies support the notion that TANs would, instead, promote tumour progression based on their ability to release angiogenic factors, and other mediators able to stimulate cell motility, migration and invasion. Almost nothing is known regarding how alteration in iron handling by neutrophils would impact on their activity in a tumoral context. These cells utilize iron-dependent ROS production through a process called respiratory or oxidative burst and iron is required for both neutrophil NADPH oxidase and MPO activities as a component of the catalytic site (34, 35). Under iron overload conditions, it has been shown that ROS production by neutrophils is significantly impaired (36). Since LUAD and LUSC infiltrating neutrophils express HEPH, making them able to avoid intracellular iron overload, this may indicate that they are fully competent to execute their cytotoxic potential, thus exerting an anti-tumour activity. They could also participate in iron clearance upon release of lactoferrin contained in their secondary granules.

In most malignancies tumor associated macrophages (TAM) are characterized by an iron-release phenotype required to sustain tumor growth and mediate immune suppression in the tumor microenvironment (37). Recently it has been shown that TAMs exposed to hemolytic red blood cells, event occurring upon extravasation of erythrocytes from newly formed vessels, can be reprogrammed from M2-like into pro-inflammatory (M1-like) phenotype, thus able to kill tumour cells (38). Interestingly these observations were performed on lung adenocarcinoma affected patients, further underlying the complexity TAM phenotypes can acquire as a consequence of different cytokine/chemokine composition found on the tumour niche.

In conclusion, our study has further underlined the complex, and still poorly understood, association existing between iron metabolism and the cancerogenic mechanisms operating in different organ landscape. Bioinformatics analysis, based on mRNA expression dataset, indicate HEPH as a novel potential prognostic biomarker for lung cancer pathologies. Up-regulation of HEPH in LUAD and LUSC correlates with a better out-come, but only from the full comprehension of the functional cross-talk occurring between the different cell types identified as HEPH-expressing cells and cancer cells will allow us to envision new therapeutic strategy to fight such devastating diseases.

## Data Availability

All data reported in the manuscript are available on request

http://ualcan.path.uab.edu

http://gepia.cancer-pku.cn

## ETHICS STATEMENT

This study was carried out in accordance with the Declaration of Helsinki and was approved by the University of Palermo Ethical Review Board (approval number 09/2018).

## AUTHOR CONTRIBUTIONS

Conception and design: Paola Zacchi, Alessandro Mangogna and Violetta Borelli. Development of methodology: Paola Zacchi, Alessandro Mangogna. Acquisition of data: Beatrice Belmonte, Letizia Scola, Gaia Morello and Anna Martorana. Analysis and interpretation of data (e.g., statistical analysis, biostatistics, and computational analysis): Alessandro Mangogna, Paola Zacchi and Violetta Borelli. Writing, review, and/or revision of the manuscript: Paola Zacchi, Alessandro Mangogna and Violetta Borelli. Study supervision: Violetta Borelli.

## ACKNOWLEDGEMENTS

We thank prof. Giuliano Zabucchi for critical review of the manuscript and Dr. Armando Gentile for his technical assistance.

